# Relationship between support for workers with illness and work functioning impairment in Japan during the COVID-19 pandemic

**DOI:** 10.1101/2021.09.23.21263920

**Authors:** Yu Igarashi, Seiichiro Tateishi, Arisa Harada, Ayako Hino, Mayumi Tsuji, Akira Ogami, Koji Mori, Ryutaro Matsugaki, Yoshihisa Fujino, for the CORoNaWork project

## Abstract

**Objective:** This study examined the relationship between job accommodations for workers with poor health and work functioning impairment during the COVID-19 pandemic.

**Methods:** An internet survey was conducted in December 2020. We included 24,429 subjects for analysis. One question was used to determine whether subjects needed job accommodations from their company to continue working in their current health condition. The odds ratios (ORs) of the necessity of job accommodations for sick workers associated with work functioning impairment were estimated using multilevel logistic regression analysis.

**Results:** The OR of work functioning impairment among sick workers not receiving job accommodations was 5.75 (95% confidence interval (CI) 5.34-6.20, p<0.001) and those receiving job accommodations was 1.88 (95% CI 1.69-2.08, p<0.001) compared to healthy workers.

**Conclusions:** This study suggests that providing job accommodations to workers with poor health may improve their work functioning impairment.

## Introduction

An estimated 80% of workers continue to work despite having an illness or health problem^1^. The development of new drugs and therapeutic devices, improved diagnostic procedures, and improved surgical techniques have led to better treatment outcomes for diseases such as cancer^2^, stroke^3^, heart disease^4^, and musculoskeletal disorders^5^, making it possible for workers who may have previously had to give up their job to instead continue working even after becoming ill^6^. In addition, extension of the retirement age has increased the number of elderly people in the workforce, who typically have a higher incidence of health problems due to their advanced age^7^.

The phenomenon of working while in poor health is known as presenteeism. The term presenteeism describes two broad concepts: a state of reduced productivity associated with poor health, and a state in which a person continues to work despite being in poor health^8^. Poor health includes having either an acute or chronic illness, and is independent of whether or not the person has been to a hospital, whether or not they have been diagnosed, and whether or not they need treatment. Workers who engage in presenteeism experience a variety of difficulties in their work. Specifically, they have a higher risk of absenteeism and unemployment^9-11^, and lower work productivity^11-13^. Moreover, presenteeism reduces workers’ quality of life^14^. In addition, prioritizing work may cause workers to miss opportunities for treatment, which may lead to severe disease^15^. To prevent the potentially detrimental effects of presenteeism on employment and the course of workers’ disease, it is important for companies to accommodate workers who desire to continue working while in poor health.

There is a growing global movement for companies to provide job accommodations to workers in poor health who want to continue to work while receiving treatment, regardless of whether they have a chronic or acute condition, such as mental illness or chronic inflammatory disease, and whether they taking anti-cancer drugs or have undergone surgery. Work accommodations include providing workers with flexible working hours so that those with poor health can work while receiving medical treatment; and employment accommodations such as job adjustments, workplace support, and physical changes in the work environment to improve their ability to work. Around the world, this concept is known as “work accommodations,” “reasonable accommodations,” or “fit note”^16-20^. In Japan, it is called “Ryoritsu Shien,” which means accommodating workers who desire to continue working despite being in poor health, with the Japanese government promoting ‘‘Ryoritsu Shien’’ to create a society where people with poor health can also work. Accordingly, government campaigns, guidelines, and a revision of medical fees have been implemented. Further, some medical institutions have established new departments to promote the accommodation of working people with poor health^21-24^. Accommodating the employment of workers with poor health has a number of benefits, including reducing loss of work productivity^25^, sickness absence^26^, the risk of job termination^16,17,27^ and job stress^16^.

It is particularly important to provide job accommodations to workers with poor health in the current COVID-19 pandemic, a period during which many have experienced interruption to treatments, particularly those for chronic, non-emergency diseases such as diabetes, chronic obstructive pulmonary disease, and hypertension^28-29^. In addition, workers with poor health suffer from psychological anxiety related to being more susceptible to COVID-19 infection and having a high risk for disease aggravation^30^, which is thought to lead to reduced work performance among those with poor health. Thus, job accommodations that make it more conducive for workers to receive treatment and provide health guidance to workers with poor health during the COVID-19 pandemic are important for maintaining and improving workers’ health condition and work performance.

We hypothesized that job accommodations for workers with poor health during the COVID-19 pandemic would improve or maintain work performance. Thus, we examined the relationship between job accommodations provided by companies for workers with poor health and a decline in work performance due to presenteeism during the COVID-19 pandemic. We also stratified our analysis by job type, because the relationship between workers with poor health and decreased work performance is hypothesized to differ by the type of job being performed. Different job types can have different characteristics, such as physical demands, loads from the physical environment, and mental strains from overwork and interpersonal communication.

## Methods

### Study design and subjects

The CORONaWork (Collaborative Online Research on Novel-coronavirus and Work) project was a cross-sectional internet monitoring survey conducted from December 22 to 26, 2020, with the aim of examining the relationship between the working environment and the health status of workers during the COVID-19 pandemic^31^. Data were collected through Cross Marketing Inc (Tokyo, Japan). From 55,045 registered monitors, 33,302 full-time workers were selected based on prefecture, occupation and sex, because we expected that random sampling without such criteria would lead to bias of participant attributes toward big cities, men, and desk workers due to the nature of internet monitoring. In addition, we sought to reduce any regional bias caused by the large regional variation in COVID-19 infection rates across Japan.

Out of a total of 33,302 participants in the survey, 27,036 were included in the study after removing those who provided fraudulent responses. Fraudulent responses were defined as follows: extremely short response time (≤6 minutes), extremely low body weight (<30 kg), extremely short height (<140 cm), inconsistent answers to similar questions throughout the survey (e.g., inconsistency to questions about marital status and living area), and wrong answers to a staged question used to identify fraudulent responses (choose the third largest number from the following five numbers). Because job accommodations such as job adjustments, workplace support, and improvements to the work environment are provided to workers by companies or supervisors, we only included employed workers. A total of 2,607 subjects who indicated that their occupation was self-employed or SOHO (Small Office, Home Office) were excluded because self-employed workers cannot receive job accommodations from the company. A total of 24,429 individuals (12,184 males and 12,245 females) were included in this analysis. This study was approved by the ethics committee of the University of Occupational and Environmental Health, Japan (reference No. R2-079). Informed consent was obtained using a form on the survey website.

### Assessment of job accommodations for workers with poor health

A single-item question was used to determine workers’ need for job accommodations: “Do you need any job accommodations from your company to continue working in your current health condition?” The participants responded by choosing from “Not necessary”, “Yes, but I am not receiving any accommodations”, and “Yes, I am receiving accommodations”. We classified subjects who answered “Not necessary” as healthy workers, “Yes, but I am not receiving any accommodations” as sick workers not receiving job accommodations from their company, and “Yes, I am receiving accommodations” as sick workers receiving job accommodations from their company.

#### Assessment of work functioning impairment

We assessed the decline in workers’ performance due to presenteeism using the work functioning impairment scale (WFun). WFun is a self-reported questionnaire developed using the Rasch model^32^. It consists of seven questions and the total score ranges from 7 to 35. A higher score indicates higher severity of work functioning impairment. A score of 21 or higher is defined as mild to severe work functioning impairment due to health problems. WFun has been confirmed for hypothesis testing, responsiveness, and criterion-relevant validity according to the consensus-based standards for the selection of health status measurement instruments. A previous study that examined the association between the interview results of occupational health nurses and WFun confirmed the validity of the criterion association^33^. In this study, we used a six-item version whose items can be equivalently converted to those of the original tool based on the Rasch model.

#### Other covariates

The following survey items were considered confounding factors: age, sex, household income, educational status, number of employees at the workplace and marriage status. Evidence from previous studies indicate that these factors are associated with presenteeism^34-37^. Although the association between these factors and job accommodations is unclear, we think that the provision of job accommodations by companies is related to socioeconomic status; this is another reason for the inclusion of the above factors as confounders in this study.

#### Statistical analysis

The odds ratios (ORs) of mild to severe work functioning impairment as assessed by WFun associated with job accommodations for workers with poor health were estimated using a multilevel logistic regression model. The model was adjusted for age, sex, household income, educational status, the number of employees at the workplace, and marriage status. We further estimated the multivariate ORs of work functioning impairment associated with job accommodations for workers with poor health stratified by job type (mainly desk work, mainly interpersonal communication, and mainly manual labor).

A p value less than .05 was considered statistically significant. All analyses were conducted using R ver.1.4.1103 (R Foundation for Statistical Computing, Vienna, Austria)^38^.

## Results

Table 1 shows the characteristics of the subjects in this study. Of the total subjects, 74.0% (n=18,120) were healthy workers, 16.0% (n=3,999) were sick workers receiving job accommodations from their company, and 9.5% (n=2,310) were sick workers not receiving job accommodations from their company. The mean age and proportion of current smokers were lower among sick workers receiving job accommodations than sick workers not receiving job accommodations from their company. In contrast, household income, educational background, and the number of employees at the workplace tended to be higher among sick workers receiving job accommodations than sick workers not receiving job accommodations from their company. The proportion of married employees was higher among healthy workers than others.

**Table 1.** Characteristic of the subjects

| Characteristic | Healthy workers<br>(n=18,120) |  | Sick workers not<br>receiving job<br>accommodations<br>from their<br>company<br>(n=3,999) |  | Sick workers<br>receiving job<br>accommodations<br>from their<br>company<br>(n=2,310) |  |
| --- | --- | --- | --- | --- | --- | --- |
|  | n | % | n | % | n | % |
| Age, mean (SD) | 46.8<br>(10.6)* |  | 45.9<br>(10.2)* |  | 44.9<br>(10.9)* |  |
| Sex, men | 9,117 | 50 | 1,985 | 50 | 1,082 | 47 |
| Household income |  |  |  |  |  |  |
| 200-299million yen | 2,256 | 12 | 770 | 19 | 340 | 15 |
| 300-499million yen | 4,216 | 23 | 1,090 | 27 | 549 | 24 |
| 500-699million yen | 3,994 | 22 | 851 | 21 | 484 | 21 |
| 700-899million yen | 3,309 | 18 | 593 | 15 | 437 | 19 |
| 900million yen or more | 4,345 | 24 | 695 | 17 | 500 | 22 |
| Education status |  |  |  |  |  |  |
| Junior high school | 211 | 1 | 71 | 2 | 26 | 1 |
| High school | 4,647 | 26 | 1,061 | 27 | 545 | 24 |
| Vocational<br>school/college, Universtiy,<br>Graduate school | 13,262 | 73 | 2,867 | 72 | 1,739 | 75 |
| Cuurent smoker | 4,504 | 25 | 1,135 | 28 | 517 | 22 |
| Number of employees at the<br>workplace |  |  |  |  |  |  |
| <10 | 2,801 | 15 | 546 | 14 | 312 | 14 |
| <100 | 5,063 | 28 | 1,179 | 29 | 621 | 27 |
| <1000 | 5,246 | 29 | 1,246 | 31 | 648 | 28 |
| >1000 | 5,010 | 28 | 1,028 | 26 | 729 | 32 |
| Marriage status, married | 10,379 | 57 | 2,034 | 51 | 1,238 | 54 |

\* Represents mean and SD.

Table 2 shows the ORs and adjusted ORs derived from multivariate models of mild to severe work functioning impairment. The odds of mild to severe work functioning impairment in age- and sex-adjusted subjects were 5.75 times higher among sick workers not receiving job accommodations and 1.88 times higher among sick workers receiving job accommodations from their company than healthy workers. After adjusting for household income, educational status, the number of employees at the workplace, and marriage status, the odds of mild to severe work functioning impairment were 5.59 times higher among sick workers not receiving job accommodations and 1.85 times higher among sick workers receiving job accommodations from their company than healthy workers.

**Table 2.**
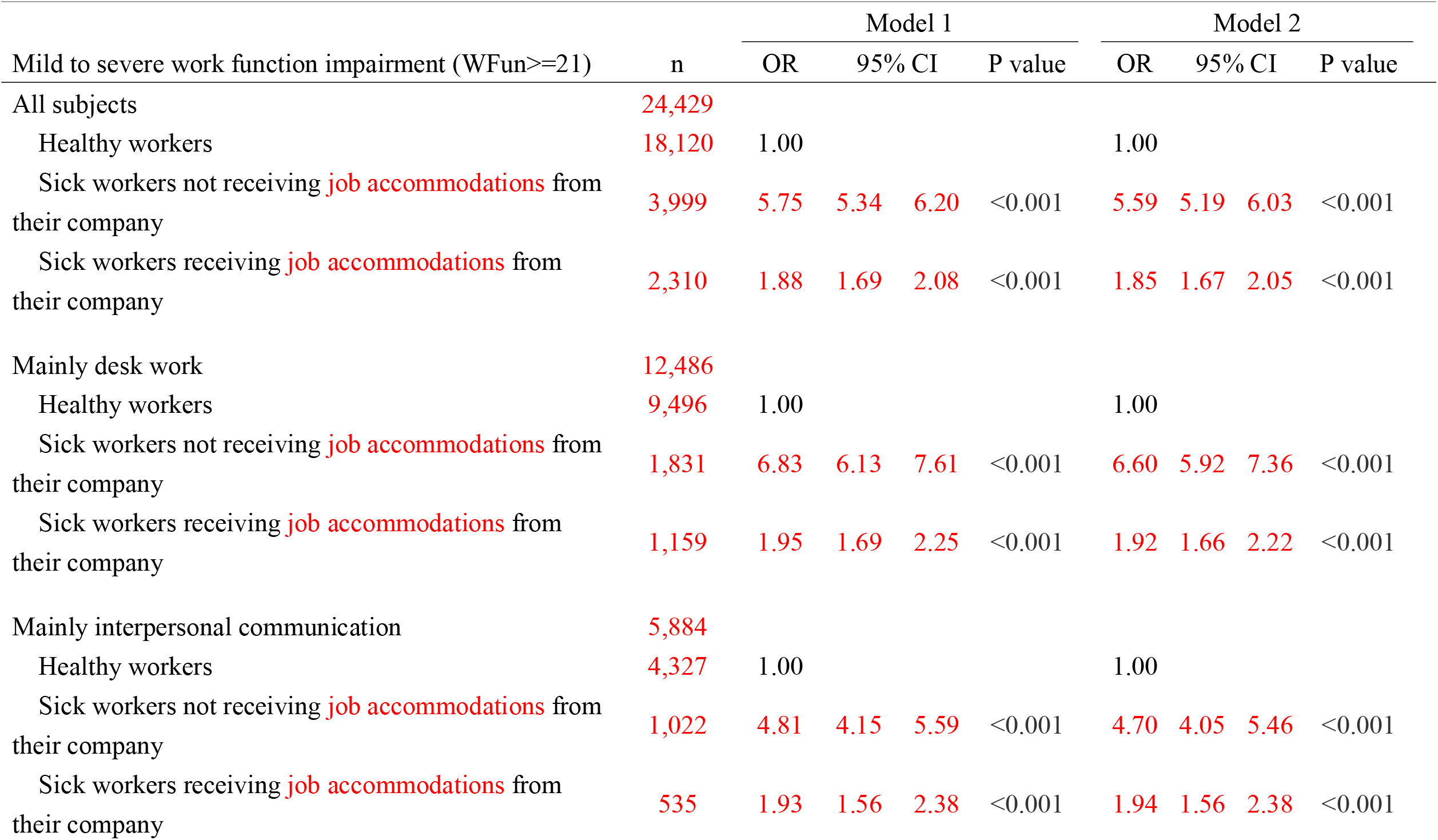
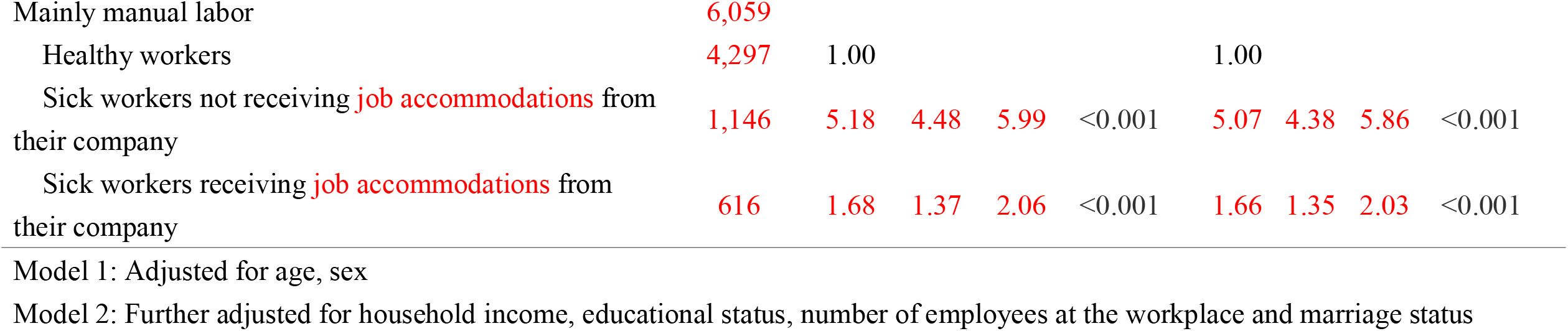
Odds ratios and adjusted odds ratios for multivariate model of mild to severe work function impairment

We also examined the relationship after stratifying by job type: mainly desk work, mainly interpersonal communication, mainly manual labor. Among subjects engaged mainly in desk work, the odds of mild to severe work functioning impairment were 6.83 times higher among sick workers not receiving job accommodations and 1.95 times higher among sick workers receiving job accommodations from their company than healthy workers. Likewise, among subjects engaged mainly in interpersonal communication, the odds of mild to severe work functioning impairment were 4.81 times higher among sick workers not receiving job accommodations and 1.93 times higher among sick workers receiving job accommodations from their company than healthy workers. Similarly, among subjects engaged mainly in manual labor, the odds of mild to severe work functioning impairment were 5.18 times higher among sick workers not receiving job accommodations and 1.68 times higher among sick workers receiving job accommodations from their company than healthy workers. Multivariate analysis showed similar trends in the adjusted odds of mild to severe work functioning impairment to those observed in the unadjusted models.

## Discussion

This study examined the relationship between job accommodations provided by companies for workers with poor health and work functioning impairment in a large population living in Japan. The odds of mild to severe work functioning impairment among workers who were not receiving job accommodations were 5.8 times higher than those of workers who did not need job accommodations. Meanwhile, the odds of mild to severe work functioning impairment among workers who were not receiving job accommodations were 1.9 times higher than those of workers who did not need job accommodations. The reduced risk of work functioning impairment for workers receiving job accommodations suggests that providing job accommodations to workers with poor health may improve worker performance. To our knowledge, this is the first study to show a relationship between job accommodations for workers with poor health and work functioning impairment in Japan.

We showed that workers with poor health have higher work functioning impairment than those who are healthy. Individuals who work with health problems have been reported to experience work disability and lower work productivity^11-13^. There are a variety of reasons why workers with poor health have reduced work ability. First, the nature of their symptoms or illness can reduce workers’ ability to perform their job or certain tasks. For example, pain or musculoskeletal diseases, which are typical causes of presenteeism, lead to decreased physical tolerance and hinder performance in tasks such as handling heavy objects. In addition, sleep disorders and mental health problems, which are a major cause of presenteeism, impair concentration and cognitive function^39^. Some conditions require workers to restrict certain tasks, such as shift work, business trips, and work at heights, to ensure safety and to avoid worsening their condition. Second, hospitalization and hospital visits for treatment may reduce working hours and interrupt work schedules. Individuals who work while experiencing work-related disability due to presenteeism have an increased risk of future days off work, job loss, and exclusion from the labor market^9-11^. Providing appropriate job accommodations to workers, such as changing the contents of their work and adjusting their working hours, is expected to decrease the loss in work performance due to presenteeism and to reduce the risk of lost work days and unemployment.

Job accommodations can reduce work functioning impairment among workers with poor health. In this study, we found that the odds of mild to severe work functioning impairment was approximately three times higher among workers who were not receiving job accommodations than those who were receiving job accommodations. Workers with poor health experience a variety of difficulties performing their duties and continuing to work. Providing job accommodations for workers with poor health in the form of adjustments to work hours and shifts, job descriptions to match their work ability, and changes to the physical work environment can help them adjust to work. Accommodations provided by supervisors and coworkers are also thought to enhance the effectiveness of these actions. Additionally, a good health climate, or the perception by employees that the team to which they belong cares about health issues, and their handling and communication of related matters, are also thought to contribute. A previous study reported that greater accommodations from supervisors and colleagues are associated with lower work productivity loss due to presenteeism^40-41^. In addition, a good health climate is associated with lower presenteeism^42-44^. Thus, job accommodations provided by companies to regulate workers’ health and work environment is expected to lead to improvements in workers’ presenteeism and performance.

We observed a similar relationship between job accommodations for workers with poor health and work functioning impairment across the job types examined. We divided occupations into three groups, namely primarily desk jobs, primarily interpersonal communication, and primarily physical labor. Different job types have different degrees of psychological and physical demands. Workers with back pain who engage in physical work naturally require different accommodations to those undergoing anti-cancer drug treatment who work at a desk. While we did not identify the details of the accommodations provided, our results nevertheless suggest that providing accommodations for workers based on their job type and health condition that are in line with their needs is useful for slowing or halting the decline in work functioning impairment across all job types. While job accommodations are increasingly being recommended for workers with poor health, there remains no unified view on the best types of accommodations to offer workers with certain illnesses and performing certain jobs. Job accommodations are currently being chosen through collaboration among workers, employers, attending physicians, and industrial physicians. In the future, studies may identify concrete recommendations of the most effective type of accommodations to provide workers according to job type.

This study had some limitations. First, because we conducted a survey of Internet monitors, a degree of selection bias was unavoidable. To minimize this, we selected subjects based on their region, occupation, and prefecture according to the cumulative infection rate from January to December 2020. Second, given the cross-sectional design, we could not determine the causal relationship between job accommodations provided by companies for workers with poor health and work functioning impairment. Workers with mild work functioning impairment who are able to work may receive job accommodations more readily. Third, we did not identify workers’ symptoms or disease in our study. The effect of job accommodations on the improvement in work functioning impairment may depend on the type and degree of poor health or symptoms exhibited by workers, combined with their job and tasks. Fourth, we used only one question to assess whether or not workers were receiving job accommodations from their company, which may not be sufficiently valid.

In conclusion, this study showed that there is a relationship between job accommodations provided by companies for workers with poor health and work functioning impairment in a large population of workers across Japan. Workers with poor health who were receiving job accommodations were less likely to have work functioning impairment than those who reported not receiving job accommodations. These results suggest that providing accommodations to workers with poor health may improve their work functioning impairment.

## Data Availability

Raw data were generated at [facility name]. Derived data supporting the findings of this study are available from the corresponding author [initials] on request.

## Acknowledgements

The current members of the CORoNaWork Project, in alphabetical order, are as follows: Dr. Yoshihisa Fujino (present chairperson of the study group), Dr. Akira Ogami, Dr. Arisa Harada, Dr. Ayako Hino, Dr. Hajime Ando, Dr. Hisashi Eguchi, Dr. Kazunori Ikegami, Dr. Kei Tokutsu, Dr. Keiji Muramatsu, Dr. Koji Mori, Dr. Kosuke Mafune, Dr. Kyoko Kitagawa, Dr. Masako Nagata, Dr. Mayumi Tsuji, Ms. Ning Liu, Dr. Rie Tanaka, Dr. Ryutaro Matsugaki, Dr. Seiichiro Tateishi, Dr. Shinya Matsuda, Dr. Tomohiro Ishimaru, and Dr. Tomohisa Nagata. All members are affiliated with the University of Occupational and Environmental Health, Japan.

